# Paucity and disparity of publicly available sex-disaggregated data for the COVID-19 epidemic hamper evidence-based decision-making

**DOI:** 10.1101/2020.04.29.20083709

**Authors:** Kristen Kocher, Arthur Délot-Vilain, D’Andre Spencer, Jonathan LoTempio, Emmanuèle C. Délot

**Author notes:** Corresponding author;, Children’s National Hospital, 111 Michigan avenue NW, Washington, D.C., USA.

## Abstract

COVID-19 has joined the long list of human disorders with sexually dimorphic expression. Increased lethality in men was evident in the first large reports from ChinaCDC and WHO-China, and the gender gap appeared even wider in the early Italian outbreak. Newspapers and scientific journals alike have commented on this finding and the preexisting conditions, biological processes, and gender role behavior differences that may underlie it. However, as for other diseases, and in spite of years of advocating for the collection of raw epidemiological data and the analysis of clinical trial data sets by sex, very little appeared to be released about sex differences in characteristics of the epidemics beyond infection and death rates, such as severity of disease, comorbidities, rate of recovery, length of hospital stay, or number of tests for the SARS-CoV-2 coronavirus. These data are critical not only for scientists to understand the pathophysiology of disease, but also to inform decision-making by countries and healthcare systems on how to prioritize testing and best allocate scarce resources and relief funds.

Systematic analysis of official websites for the 20 countries and 6 US states reporting the highest number of cases on March 21, 2020, revealed a wide disparity in sex-disaggregated data made available to the public and scholars. Only a handful of the countries reported cases by sex separately. None of the other characteristics, including fatality rates, were stratified by sex at the time. Beyond suboptimal sex disaggregation, our analysis found a paucity of usable raw data sets and a generalized lack of standardization of captured data, making comparisons difficult. A second round of data capture in April found more complete, but even more disparate, information.

Our analysis revealed a wide range of sex ratios among confirmed cases, which changed over time. In countries where a male-biased sex ratio was initially reported, the reported proportion of women among cases dramatically increased in under 3 weeks. In contrast, men were consistently over-represented in severe cases, intensive care admissions, and deaths. We also show that the sex ratio varies with age, with a complex pattern, reproduced across the 6 countries for which data were found.

Accurate, peer-reviewed, statistical analysis of harmonized, sex-disaggregated data for other characteristics of epidemics, such as availability of testing, suspected source of infection, or comorbidities will be critical to understand where the observed disparities come from and to generate evidence-based recommendations for decision-making by institutions and governments around the world.

---

In February 2020, two early reports on the epidemiological characteristics of the COVID-19 outbreak on large populations were made public, both from China. The first, published by the China Center for Disease Control (CCDC) on February 17, 2020^1^ included 72,314 individuals suspected of being infected, of which 44,672 (61.8%) were confirmed by laboratory testing. Among those, it reported a (male-to-female) sex ratio among cases of 1.06:1 (with slight geographical variation, 0.99:1 in Wuhan and 1.04:1 in Hubei). The second report, published less than 2 weeks later by the World Health Organization (WHO)-China Joint Mission on COVID-19^2^, confirmed that roughly an equal number of men and women were infected in this population (51.1% of 55,924 laboratory-confirmed cases were men). Both studies also calculated a case fatality (or lethality) rate, and found a large sexual dimorphism. The earlier study found that 2.8% of infected men vs. 1.7% of women had died. The later report had higher lethality rates but a similar ratio (crude fatality rate was 4.7% in men vs. 2.8% in women). For a roughly similar infection rate, the distribution of lethal cases was almost 2/3 of males and 1/3 females, meaning twice as many men died of the disease than women.

A New York Times article on March 20, 2020^3^ described a similar sexual dimorphism for the Italian outbreak: according to an analysis by the Higher Health Institute of Rome of over 25,000 COVID-19 cases, 8% of infected men, compared to 5% of infected women, had died. Possible epidemiological differences also began to emerge as, in the Italian population, the proportion of men in the group who tested positive was significantly higher (about 60% were men) than in the Chinese cohorts.

This finding was reminiscent of the two previous outbreaks due to other strains of coronavirus, Severe Acute Respiratory Syndrome coronavirus (SARS-CoV) and Middle East Respiratory Syndrome coronavirus (MERS-CoV). For example, the lethality in infected men during the 2003 SARS-CoV outbreak in Hong-Kong was 21.9% vs. 13.2% in women^4^; lethality has also been higher in men in the MERS-CoV outbreak (e.g. 30.5% vs. 25.8% in 2017^5^). Possible explanations for the sex differences in SARS-CoV-2 infection have been extensively discussed elsewhere^6–8^ and include sexual dimorphism in immune response to infection, incidence of comorbidities and risk factors, such as chronic lung disease, and the gender-role behaviors that may underlie those, such as smoking.

While the United Nations have requested gendered statistics be collected since at least the 1975 first World Conference on Women, a December 2010 report from the UN Economic and Social Council on the Global Gender Statistics Program highlighted that many countries in did not in many domains^9^. This was also true for health-related data, as documented during the third Global Forum on Gender Statistics (Manila, 2010; which focused on health statistics). Stunningly, it took the WHO until 2019 to publish its annual World Health Statistics report with data disaggregated by sex^10^. This report highlighted the vast sex differences in all aspects of health epidemiology, (including access to and attitudes toward care), and revealed that “of the 40 leading causes of death, 33 causes contribute more to reduced life expectancy in men than in women”. The importance of, and suggestions for, metrics to document communicable diseases were also detailed in a 2007 WHO report^11^.

Recent experiences with the Ebola virus and Zika virus epidemics should have educated us to the importance of documenting epidemiological metrics by sex, adapting aid response to the specificity of local gender roles, and involving both genders in surveillance and decision-making^12^. However, as Wenham et al. alerted in The Lancet on March 6, 2020^12^: “Policies and public health efforts have not addressed the gendered impacts of disease outbreaks. The response to coronavirus disease 2019 (COVID-19) appears no different. We are not aware of any gender analysis of the outbreak by global health institutions or governments in affected countries or in preparedness phases.”

Mandates to address sex/gender in grant applications have been in place since 2010 in Canada^13^ and 2014 in Europe^14^. Inclusion of women in NIH-sponsored clinical trials has been required since 1993 and, since 2015, NIH policies requires consideration of the concept of *sex* as *a biological variable (SABV)* in the design, analysis, and reporting of studies^15^. Guidelines to design such studies have been published (e.g.^16^). However, while researchers are now routinely reporting sex ratios in their cohorts, the full potential of analyzing sex-disaggregated data to uncover potential sexual dimorphisms has yet to be realized. For instance, the first COVID-19 randomized clinical trial, reported that lopinavir-rotonavir, an antiviral combination used in the treatment of HIV infection, was not effective against SARS-CoV-2^17^. A similar number of men and women were included in the trial, but drug efficacy was not analyzed by sex. Similarly, a report demonstrating that heart damage is in independent risk factor for death in COVID-19 infection did not analyze data with sex as a biological variable^18^. A similar number of men and women (50.7%) were included in the final report of 416 consecutive cases, but a larger proportion of women had been excluded (57% of 229) because critical laboratory results were not available for them. This seems a missed opportunity given the well-known sex differences in the burden of cardiovascular disease (e.g.^19^).

An emerging hypothesis to explain the severity of disease in some patients is the so-called “Cytokine Storm”, or cytokine release syndrome^20,21^. Accounts are emerging in the news media that critical patients were saved by experimental treatment with anti-inflammatory drugs (e.g.^22,23^). As the immune response is one of best documented sexually dimorphic biological process in humans^24,25^, collecting and analyzing data regarding the immune and inflammatory status of patients in a sex-disaggregated way will be critical in devising appropriate therapies.

We sought to document what sex-disaggregated information was made available to the public and the research community through governmental websites. To account for the extreme fluidity of the situation, we accessed official websites at the transnational, governmental, and US State levels in on two occasions within the span of approximately 3 weeks, between March 21-24 and April 5-10, 2020. Available sex data for cases and deaths were scarce in the first data capture, and more generalized in the later one. We found extreme diversity in the type of data captured, graphs chosen to illustrate them, and ease of accessibility of released information. This diversity made it very difficult to compare data from country to country and to derive conclusive lessons from the data currently available. The abundance of non-standardized information illustrates both the nimbleness of digital systems to support responsiveness to a new pandemic and the lack of preparedness of governmental health authorities worldwide for such an event.

## METHODS

On March 22, 2020, we accessed the list of world countries ranked by number of reported cases of COVID-19 maintained by Worldometers.org. Between March 22 and March 24, we accessed the sources quoted by Worldometers to search for raw data. These sources included national news outlets reporting about official press releases (*e.g*. El País for Spain), as well as official governmental websites. When no governmental website was quoted, we independently searched for one. We visited and took timed-stamped screenshots of all the websites. Websites were searched by native speakers in English, French, Chinese, and Swedish; authors with fluent or functional knowledge of the language in Spanish, Italian, and German, further aided by online translators for Portuguese, Dutch, Norwegian and Danish. Available metrics, source URLs, and date of accession are collated in **Table 1**.

**Table 1:**
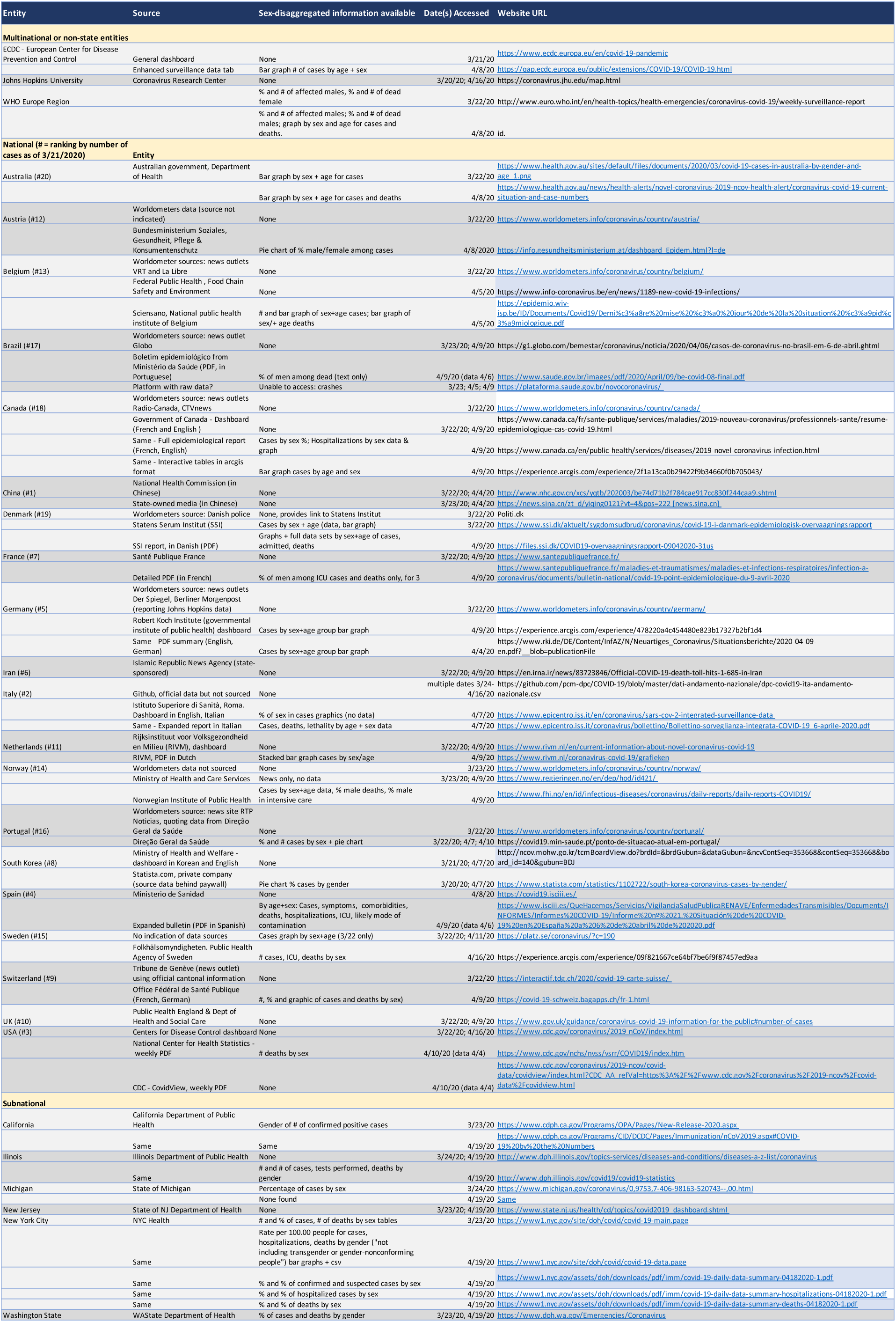
Sex-disaggregated metrics documenting the COVID-19 epidemic available on official websites. Data are shown for the 20 countries with the highest number of reported cases on March 21, 2020 (ranking shown in parenthesis in column 1), the six US states and 3 trans-national or non state entities. Data source is described in column 2. Metrics found on the websites indicated in column 5 are indicated in column 3. Dates of accession are shown in column 4. If a website showed different metrics at a different capture date, these are indicated in separate lines. If different information was shown in different format (e.g. dashboard vs. pdf), URLs are shown separately.

We also accessed the websites of trans-national entities, such as the WHO and the European CDC. Deficiencies of the WHO website, including a change in reporting methods on March 18 and an inability to correct errors in a timely fashion, were documented by ourworldindata.org^26^. The ECDC, which reports daily statistics for the whole world, beyond Europe, has been used as a reliable source by other reputed outlets. Johns Hopkins University also maintains a tally of cases on its Coronavirus Report Center webpage used as a source by news outlets. We were unable to locate any sex-disaggregated metrics on the ECDC (accessed March 20, March 22, April 2), Johns Hopkins (March 20, April 2), WHO (March 21; April 12), Worldometers (daily March 20-24), or the CDC (March 20, April 2) dashboards.

As the US CDC disclaimed that they had stopped tallying tests and recovery rates, deferring to each US State’s authorities to do so, we also sought data provided by the six States with the highest number of cases on March 23-24, 2020: New York, New Jersey, Washington, California, Michigan, and Illinois.

As this first search revealed large variability in types of data collected and reported by governmental sites, as well as in sexual dimorphism trends, we performed a second data capture to detail the diversity of reported metrics. Websites were re-accessed at a later date, between April 4 and 10, 2020, where possible by more than one of the authors, to verify and complete data sets. (Details of reported metrics not related to sex will be reported elsewhere^27^.)

To verify whether the trend of longitudinal sex ratio decrease among confirmed cases identified in some countries (see below) held in US States, the same 6 state websites were re-accessed on April 19 (by which time New York City had more confirmed cases than any world country).

## RESULTS

### Reported metrics in early (Mar 21–24) data capture

Worldometers did not provide sex-disaggregated statistics. It cited governmental websites for 8 countries, news agencies for 8, and no source for 2 (**Table 1**). For the remaining two, websites with uncredited sources were quoted: one is a GitHub for the (presumably official) Italian data; for the other, on platz.se, for Sweden, we could not identify the authors or from where the data is sourced.

We found documentation of the sex ratio among confirmed cases on the main governmental interface and/or the Worldometers source for 6 of 20 countries, 4 of 6 US states and the WHO-Europe website. Italian data was reported in the New York Times^3^. Sweden and Australia provided graphs of cases disaggregated by both sex and age (but no numerical values). Only Denmark provided both a graph and the attached numbers. For China, available information was from the published reports mentioned above^1,2^; we were unable to find sex-disaggregated data on the interface of either the ChinaCDC or the state-sponsored news agency SINA (which both showed the same data when simultaneously accessed on 4/4/20). We were able to find a sex ratio of deaths for 2 countries (Italy and South Korea) in the news media, and on the websites of 2 US entities (Washington State and New York City), and the Europe-WHO region.

Several countries reported admission to Intensive Care Unit as a proxy of severity of disease (*e.g*. Italy, Belgium), but none did so in a sex-disaggregated way. No other metrics of the COVID-19 epidemic shown on various dashboards, such as regional distribution, possible source of infection, Intensive Care Unit (ICU) hospitalization, number of tests performed, recovery rate, or comorbidities, was reported by sex.

### Reported metrics in the second (April) data capture

By the time of our second data capture in early April 2020, most, but not all, countries had started reporting cases by sex (typically percentage of confirmed cases; **Table 2**). For some countries, such as the US or France, the sex-disaggregated data was not provided on the main public dashboard, but searching through layers of links located more specific reports where the information could be found.

**Table 2:** Diversity and evolution of reported sex ratios for COVID-19 infection. Sexual dimorphism of COVID-19 infection rates is shown for the only countries and US States for which data were available on the websites of the 20 countries and 6 US states with the highest number of cases as of March 21, 2020 (March column). Trends and numbers are shown, when available, for the April capture (Apr. 7–9 for countries; Apr. 19 for US States, lilac cells). **Countries reporting over 10 percentage points more of men or women are shown in blue and red respectively**. Where higher proportions of infected men were reported in the early data capture, the proportion of women had dramatically increased by the time of the second data capture. *China-WHO data from the Feb. 28 report (ref. 2). **Italy: Higher Health institute of Rome data, quoted in the NY Times (ref. 3). ***WA state data published as 51% female, 45% male, 4% unknown sex for 2,221 cases. Shown is calculated rate for cases where sex is known (1,133 F/999 M).

| Confirmed cases | Proportion of women |  | Trend | Number of cases in report |  |
| --- | --- | --- | --- | --- | --- |
| Capture date | March | April |  | March | April |
| China WHO * | 48.6% | no data | ? | 55,924 | n/a |
| Denmark | 39.3% | 55% | ↑ | 1,395 | 5,635 |
| Italy** | <40% | 47.1% | ↑ | 2,5058 | 124,162 |
| Portugal | 51.3% | 56.7% | ↑ | 8,799 | 12,442 |
| South Korea | 61.5% | 60% | ↔ | 1,600 | 10,284 |
| California | 43.8 % | 49.4% | ↑ | 2,102 | 28,720 |
| Michigan | 48% | no data | ? | 1,791 | n/a |
| New York City | 43% | 47% | ↑ | 12,339 | 126,098 |
| Washington State *** | 53 % | 53 % | ↔ | 2,132 | 11,448 |

#### Cases

We found sex data for confirmed cases in graph-only format for 3 countries (Australia, Germany, Netherlands), and numerical values for another 11 countries. For another, France, we were able to find sex-disaggregated numbers only for ICU cases. Brazil provided numbers and a bar graph disaggregated both by sex and age, but with all cases of Severe Acute Respiratory Distress Syndrome aggregated together. These included Influenza A, COVID-19 and, for the vast majority, cases still under investigation or undiagnosed.

#### Deaths

Sex information for deaths was found for 12 of the 20 countries: Australia and Belgium in graph-only format and numerical data for Brazil, Canada, Denmark, France, Italy, Norway, Spain, Sweden, Switzerland, the USA (CDC, IL, WA, NYC).

Sites with already detailed information in March, deployed richer data and analyses 2–3 weeks later. For example, Denmark, the first country to provide numerical data for cases disaggregated both by sex and age in March, provided both graph and numerical data disaggregated by sex, age, and comorbidity for cases, deaths, and hospitalizations in April (in PDF format, in Danish).

#### Comorbidities

The first (and only) country to report comorbidities (and symptoms) disaggregated by sex was Spain.

#### Tests

The first (and only) report of sex information on the number of tests performed was found April 19 for the US state, Illinois.

#### Other

At the time of writing, no information had been found by sex for suspected source of infection or recovery rate.

### Variability of sex ratio among confirmed COVID-19 cases by country (early data)

The February reports from China indicated a similar number of men and women among confirmed cases (51.1% of men of almost 56,000 cases^2^). However soon South Korean data emerged that looked very different, with a large excess of women (61.5%) among 8,799 confirmed cases^28,29^. The data from Italy around the same time showed an equally strong sex bias, but in an inverse direction, with over 60% of infections in men out of over 25,000 cases^3^. [Information came in from either a press article, the Tweeter feed of a reputed sex differences researcher, or the analysis of a private company]. The only other two countries for which we found numerical data in March, Denmark and Portugal, failed to bring any clarity: the sex ratio in Denmark was similar to that of Italy, Portugal’s closer to China’s with 48.7% of men. In US states, the sex ratio ranged from 43% of men in Washington State to 56.2% in California (all data are shown in **Table 2**).

### Variability of sex ratio among confirmed COVID-19 cases by country (April data)

More countries reported cases by sex in the later round of data capture, but the disparity of sex ratio persisted. The format in which those were reported was also variable: number of cases, percentage of men, and/or pie charts. Pie charts were visually impactful for data comparison; they are shown in **Fig. 1** for the countries that made them available. All available numbers are compiled in **Table 3** (where values provided by the websites are highlighted in green cells, while values calculated by the authors are indicated in italics).

**Fig. 1.**
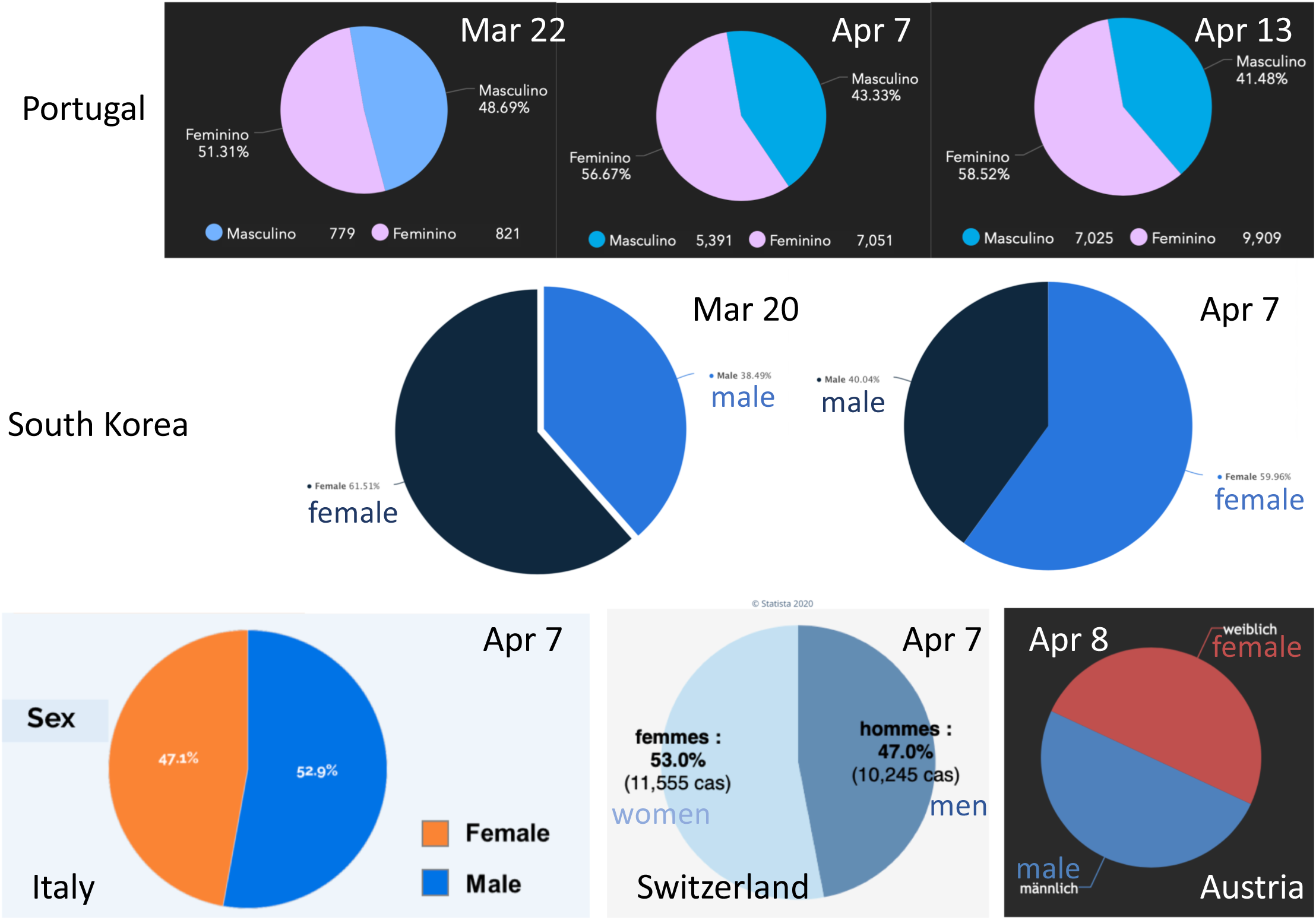
Visual comparison of sex ratio among cases by pie charts. Screen shots of pie charts found for 2 and 5 countries in March and April respectively. Longitudinal comparison in Portugal shows an increase in the percentage of women among cases. In the same time period proportions have been roughly unchanged in South Korea (with ~60% women), although the change of color convention of this statista.com graph is misleading. Sex ratios are inverse in Italy (52.9% men) and Switzerland (53% women), while equal numbers of affected men and women were reported in Austria. (Text in English in appropriate color was overlaid over the screen shot when needed for increased legibility). Date of data is indicated.

**Table 3:** Availability of sex-disaggregated data captured April 5–10, 2020 for cases and deaths on the official websites of the 20 countries with the highest numbers of cases as of March 21, 2020. Cells highlighted in green represent the metrics actually shown on the website (dashboard or PDF). Numbers in italics have been calculated by the authors for this publication. In bold is the proportion highest of the two sexes. * Sex-disaggregated data were available for only a subset of the cases known on that day. It represented 70% (of 19,774) for Canada. For Spain, on the date of data capture (4/9), sex-disaggregated data was available only for cases up to 4/6, and represent 54.9% of the known cases (152,162) and 39% of deaths (15,238) on capture day. Italy reported 365 cases with sex unknown, and data shown represent 99.7% of known cases. ** The CDC dashboard, accessed April 10, reported cases for the 1 Mar - 4 Apr period and deaths for the 1 Feb-4 Apr period. No sex-disaggregated data could be found for cases. *** Sweden data was obtained at a later date, from another, governmental website, as the website originally accessed had ceased reporting data by sex and did not identify their data source.

|  |  | CASES |  |  |  |  | DEATHS |  |  |  |  |
| --- | --- | --- | --- | --- | --- | --- | --- | --- | --- | --- | --- |
|  | Date for sex data | Number in report with known sex | Number of women | Number of men | % of women | % of men | Number in report with known sex | Number of women | Number of men | % of women | % of men |
| Australia | April 5 | 5795 | graph only | graph only | graph only | graph only | 39 | graph only | graph only | graph only | graph only |
| Austria | April 8 | 12,942 | 6,471 | 6,471 | 50 | 50 | no data | no data | no data | no data | no data |
| Brazil | April 9 | 15,927 | no data | no data | no data | no data | 655 | 268 | 387 | 40.9 | 59.1 |
| Belgium | April 5 | 19,589 | 10,681 | 8908 | 54.5 | 45.5 | 1477 | graph only | graph only | graph only | graph only |
| Canada | April 9 | 14,167* | 7,508 | 6,659 | 53 | 47 | 461 | no data | no data | no data | no data |
| China WHO | April 10 | 83,305 | no data | no data | no data | no data | 3,345 | no data | no data | no data | no data |
| Denmark | April 9 | 5,635 | 3,076 | 2,559 | 55 | 45 | 237 | 92 | 145 | 38.8 | 61.2 |
| France | April 10 | 90,676 | no data | no data | no data | no data | 13,197 | no data | no data | no data | no data |
| France (death certificates) | 1 Mar-6 Apr | n/a | n/a | n/a | n/a | n/a | 3,975 | 1681 | 2,294 | 42.3 | 57.7 |
| Germany | April 9 | 108,202 | graph only | graph only | graph only | graph only | 2,107 | no data | no data | no data | no data |
| Iran | April 9 | 66,220 | no data | no data | no data | no data | 4,110 | no data | no data | no data | no data |
| Italy | April 7 | 124,162* | 58,172 | 65,990 | 46.9 | 53.1 | 14,840 | 4,793 | 10,047 | 32.3 | 67.7 |
| Netherlands | April 9 | 21,762 | graph only | graph only | graph only | graph only | 2,396 | no data | no data | no data | no data |
| Norway | April 9 | 6,160 | 3,082 | 3,078 | 50 | 50 | 88 | 40 | 48 | 45 | 55 |
| Portugal | April 7 | 12,442 | 7,051 | 5,391 | 56.7 | 43.3 | 345 | no data | no data | no data | no data |
| South Korea | April 7 | 10,284 | 6,166 | 4,118 | 60 | 40 | 186 | no data | no data | no data | no data |
| Spain | April 6 | 83,633* | 42,448 | 41,185 | 50.8 | 49.2 | 4,409 | 1,615 | 2,794 | 36.7 | 63.3 |
| Sweden | April 15*** | 11,927 | 6,143 | 5,784 | 51.5 | 48.5 | 1,203 | 517 | 686 | 43 | 57 |
| Switzerland | April 9 | 24,140 | 12,876 | 11,264 | 53.3 | 46.7 | 804 | 311 | 493 | 38.7 | 61.3 |
| UK | April 9 | 65,077 | no data | no data | no data | no data | 7,979 | no data | no data | no data | no data |
| USA CDC | April 10 ** | 22,601 | no data | no data | no data | no data | 4,065 | 1,648 | 2,417 | 40.5 | 59.5 |
| Europe WHO Region | April 8 | 95,401 | 45,660 | 49,741 | 48 | 52 | 8,569 | 2,666 | 5,903 | 31 | 69 |

We were unable to find the number of total cases disaggregated by sex for 6 countries (Brazil, China, France, Iran, UK and USA). Three countries reported cases by sex only in graph form (Australia, Germany, Netherlands), disaggregated by both sex and age. Visual inspection of the graphs suggested a large surplus of men among cases in the Netherlands, but not in Germany or Australia (see **Fig. 2**). Among the 11 countries where numbers were available (**Table 3**), the proportion of men varied by over 13 percentage points, ranging from 53.1% (Italy) to 40% (South Korea). Italy was the only country with a larger proportion of men. Two countries reported an equal number of men and women among confirmed cases (Austria and Norway; with Spain at 50.8%). Belgium, Canada, Denmark, Portugal, S. Korea, and Switzerland reported an excedent of women by at least 6 percentage points.

**Fig. 2:**
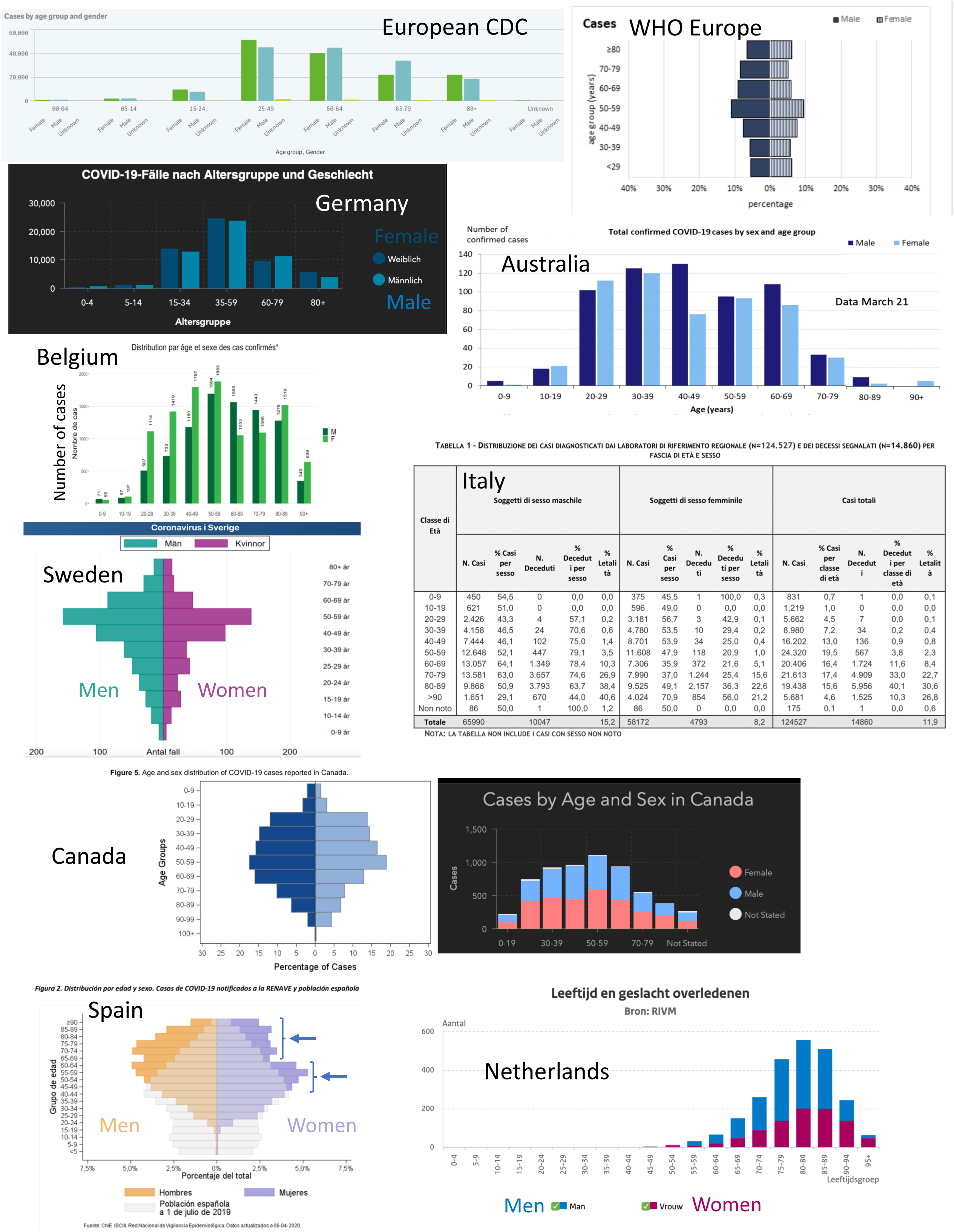
Wide disparity of data display and age binning for COVID-19 cases disaggregated by sex and age. Screen captures of data disaggregated by both sex and age were found for 9 of the 20 countries (Denmark is shown in Fig. 2) and Europe. ‘Men’ and ‘women’ labels have been added to the screenshots, in the appropriate colors, when not in English. Blue arrows on the Spain graph were added to indicate features (see description in text). Age-binning, reported metric, and type of graphic representation was different for all.

### Increase in the percentage of women among confirmed cases between the two data captures

The pie chart representation of the Portuguese data helped us immediately visualize a change of sex ratio among cases between March 22 and April 6. As shown in **Fig. 1** (top row), longitudinal comparison in Portugal revealed an increase in the percentage of women among cases from 51.3% (Mar 22) to 56.7% (Apr 7). The trend continued with an increase to 58.5% just 6 days later.

In the same time period, proportions appeared roughly unchanged in South Korea, with ~60% women, (although the change of color convention of the statista.com graph misleadingly suggests otherwise; Fig. 1 middle row). Among the other countries who had started providing pie charts in the second capture (Fig. 1 bottom row), Austria had equal numbers of confirmed infected men and women, and Switzerland more women (53%). In Italy, which had reported a large excess of infected men earlier (60%), the percentage had now fallen to roughly 53%.

To verify if the trend held, we also recaptured data for the 6 US states (on April 19, lavender cells in **Table 2**). Data by sex could no longer be found on the Michigan website. Washington still reported the same higher proportion of female cases (53%) as before. However, in both New York City and California, which had a greater percentage of confirmed male cases in March (~57%), the percentage of infected women had increased (to 47% and 49.4% respectively).

To further understand the trend, we turned to the reports of data disaggregated by both sex and age. On March 22, the only country providing numerical values for this was Denmark. **Fig. 3** shows the evolution of the data from Denmark in the captured mirror bar graphs. While the data showed many more infected men than women in all age groups on March 22, the trend had inverted by April 8, with many more infected women than men between the ages of 20 and 60 (confirmed cases increased almost 8-fold in that timespan).

**Fig. 3.**
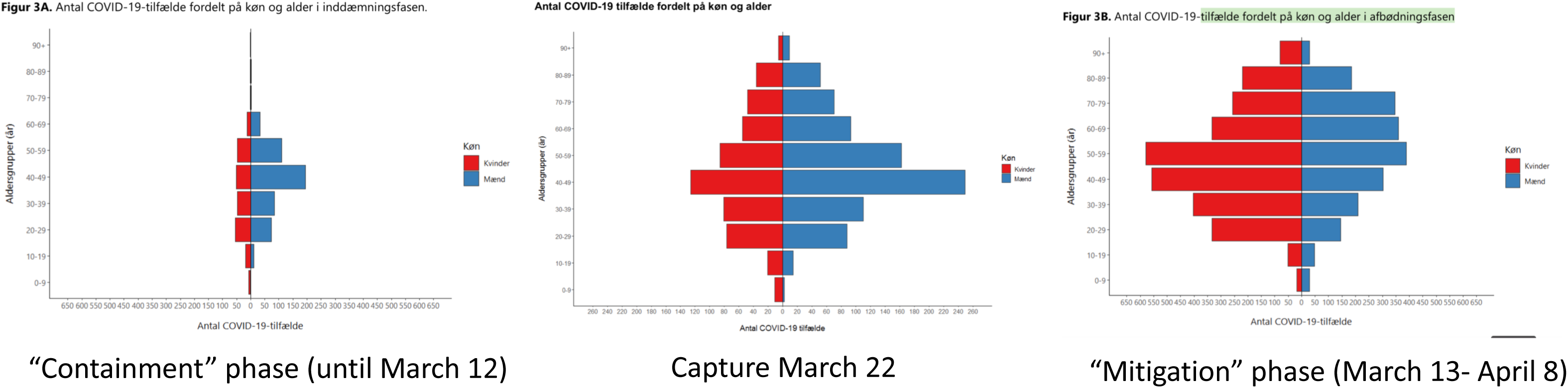
Mirror bar graphs of the number of confirmed cases by age. [Y-axis: age in 10-year bins] **and sex** [X-axis: number of cases, scale 0–650 (left, right) or 0–260 (center)] **in Denmark illustrate widely different distributions over time**. The three graphs illustrate the difficulty faced by scholars analyzing data in real time to try and derive evidence-based recommendations. The left (cumulative cases up to March 12) and right (cumulative cases March 13-April 8) graphs were captured on April 9 on the same website as the March 22 capture (center). Red: women; blue: men. (Numbers were also provided in accompanying tables).

### Disaggregation of confirmed cases by both sex and age

While most countries provided disaggregated data by age, disaggregation by both sex and age was rare in the first data capture. That information, while likely available, was also not included in the reports on Chinese data^1,2^.

As of March 24, we had been able to find such information for only three countries. Two, Sweden and Australia, showed a graph without attached numerical values - one a bar graph, the other a pyramid graph, using different age categories (**Fig. 2**). Denmark showed a pyramid graph (**Fig. 3**) and provided a table with the numbers used to create it. Both Sweden and Denmark had clearly more male cases than female at all ages. In the Australian cohort, counts were similar across age groups, except in the 40-49 age range, where women represented only about 40% of the cases.

In the second round of data capture, we were able to find information disaggregated by both age and sex for 9 of the 20 countries and Europe, typically in analytical PDFs linked from the main dashboard. For example, a bar graph, with indicated numbers, can now be found in the rich PDF, updated daily by the Sciensano Institute in Belgium, available only in French. (The dashboard of the Belgian Federal Public Health, visible in French, English, Dutch and German, has no gendered information). The Istituto Superiore di Sanità in Rome, Italy, also publishes a daily-updated table with cases and deaths by sex and age (available in Italian, in PDF format, as a link off the Italian or English interfaces of the Epicentro website). Switzerland displays the data on a highly interactive dashboard with cases, deaths numbers, percentages and clickable illustrations (in French and German).

While more information was made available between the first and second data capture, the extreme diversity of representation and metrics made comparing data between countries difficult, as illustrated in **Fig. 2**. The EuroCDC displayed the number of cases in Europe in side-by-side bar graphs for females and males, accounting for missing data (“unknown”), while WHO Europe displayed the percentage each age group represents among cases in males and females in mirror bar graphs using an oddly expanded X-axis scale. Age was binned in 5-, 10-, 15- or 25-year increments across the lifespan by EuroCDC and by grouping ages 0–29 years, then in 10-year bins by WHO Europe. A graph for Germany was similar to that of EuroCDC, except it showed only 6, different age groups (0–4, 5–14, 15–34, 35–59, 60–79, 80+) in irregular (5-, 10-, 20 - or 25-year) increments. Two other countries also displaying numbers of cases by sex in side-by-side bar graphs used regular 10-year age bins (Australia and Belgium; in addition to the graph, numbers were provided for Belgium but not Australia).

No graph was provided for the Italian dataset but a very complete table included, in bins of 10 years, case (and death) numbers for each sex, percentage of each sex among cases (and deaths), and total (including those cases where sex was not documented).

Two linked websites illustrated the Canadian data with different metrics and graphs: mirror bar graph of case percentages by age in ten 10-year bins (plotted age 100 to 0, opposite to all other graphs) (www.canada.ca) vs. stacked bar graphs of number of cases (https://experience.arcgis.com). The former did not have a legend for the color by sex on the graph (when we returned 5 days later to verify the data, the URL had become inactive). For the latter, the X axis indicates 4 non-continuous age groups (+ unknown) but 9 bars are shown, and it is unclear what age ranges are actually shown (or if this is browser- or screen-size-specific). One other country showed the number of cases by sex in a stacked bar graph, the Netherlands, but used a different age binning (with 20 categories of regular 5-year increments).

Finally, mirror bar graphs (pyramids) were chosen to display data from Sweden (on March 22 but no longer available in April), Spain (available in April, but not yet in March) and Denmark. Sweden and Denmark showed number of cases, while Spain plotted percentage of each age bin among the cases. Age was binned differently by the three countries: increments of 10 years for Sweden, except between ages 10–30, which were split into 5-year bins; 19 groups of 5 years for Spain; 10 groups of regular 10-year bins for Denmark. Denmark and Spain also provided numerical data in attached tables.

Spain helpfully overlaid a pyramid of ages of the general population, which clearly illustrates the low rate of infection in people under the age of 20 and the high rate among men above age 50. Interestingly, for women, it suggests a bimodal effect depending on age (starting around the time of menopause and above age 60; blue arrows in Fig. 3).

### Sex ratios vary by age in a consistent pattern across countries

**Figure 3** illustrates the difficulty of comparing results when representations, age binning, and metrics (number vs. percentage of each age group among cases) are not standardized, and no clear trends were immediately apparent. We used the numerical data made available by Belgium, Denmark, Italy, Norway, Switzerland, and Spain to plot the sex ratio across the ages (**Fig. 4**; numerical data and data sources are shown in **Supplementary Table 1**). Both the datasets from March 22 and April 9 for Denmark are represented.

**Fig. 4.**
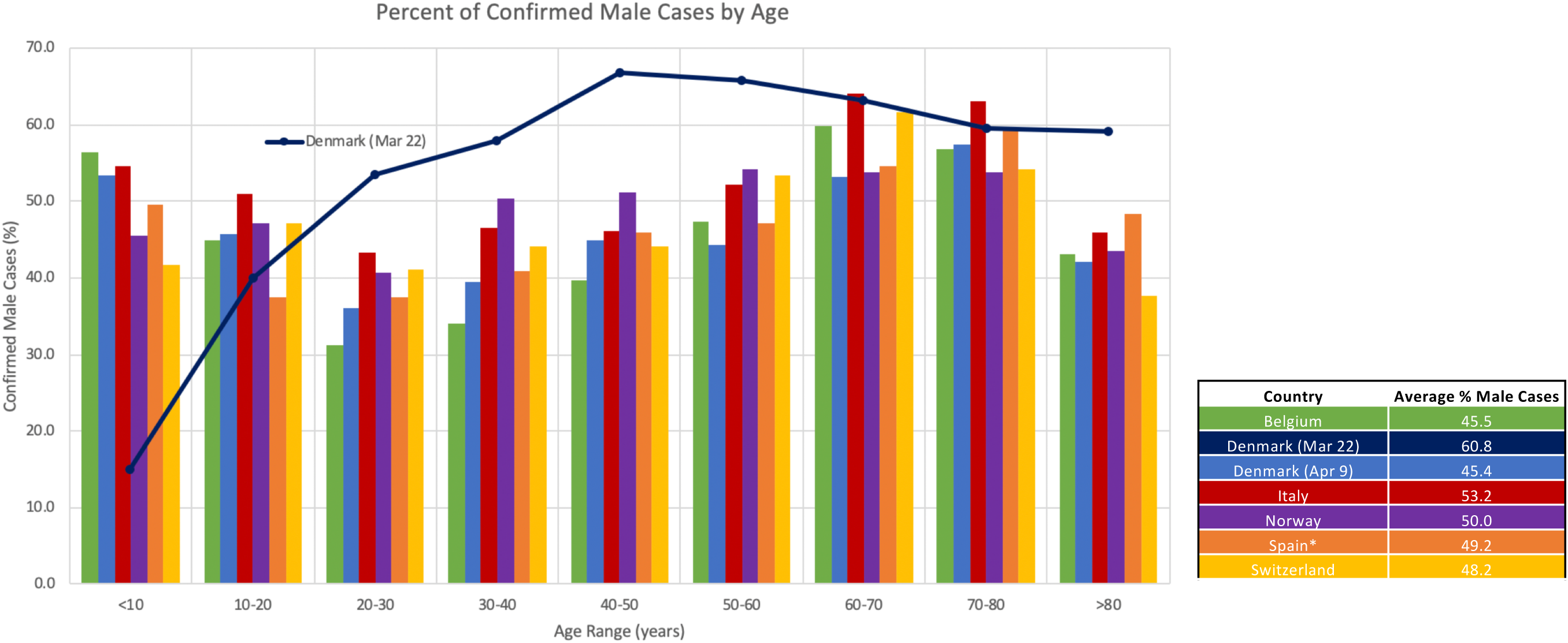
The sex ratio of confirmed infections varies by age and with time. Proportion of men by age, binned in 10-year increments, is shown. Data for Denmark are graphed at 2 stages of the epidemics: Mar. 22, when it was the only such data available (dark blue line), and Apr. 9 (medium blue bars), when the sex ratio at all ages mirrored trends in the other countries captured at a similar time. The average percentage of men across all age groups for each country is shown in the inset table. All data, data source, and calculations are shown in Supplemental Table S1. Captures were on Apr. 5 for Belgium, Apr. 7 for Switzerland and Italy (captured data also shown in Fig. 3), Apr. 9 for Spain (data 4/6) and Norway.

This revealed that the sex ratio among cases varied with age following a complex trend. For all 6 countries, the sex ratio decreased from birth to a low at age 20–30 (when ~2/3 of cases were in women in Belgium). It then increased up to age 60, plateaued until age 80, when it started decreasing again (likely due in part to the excess of women in the general population in that age range). This trend was seen in all six countries, in spite of their very divergent average sex ratios [range 45 to 53%].

Strikingly, the sex ratios calculated for the Denmark April 9 data followed the same trend as the other 5 countries, in sharp contrast to the monophasic trend observed for the March 22 data (**Fig. 4**).

### Men accounted for the majority of casualties of COVID-19 in all countries

#### Early data

As of March 24, sex information about deaths had been made available for 3 countries: Italy (data 3/24; NY Times), South Korea (3/21; Dr. Klein’s Twitter feed^29^), and China (February^1,2^). We did not find sex-disaggregated data for casualties on any of the websites for the 20 countries. We did find this information on the websites of WHO-Europe region, Washington State, and New York City.

China had reported a much higher lethality (deaths among confirmed cases, also known as case fatality) of 4.8% in men vs. 2.8% in women (~36% of casualties were women^2^). WHO-Europe reported 28.6% of females among the 1,032 deaths during the week of March 9–15, very similar to the 29% reported for Italy (cumulative to March 20). New York City seemed to concur with 31.8% of women (among what was still a low number of deaths, 63). In contrast, South Korea and Washington State reported much higher proportions of women: 47% (of 102 deaths) and 55% (of 108), respectively. As indicated above both had much higher proportion of infected women as well, even early into the epidemic.

#### Confirmation of trend

At the second round of data capture, we were still unable to find deaths stratified by sex data for Austria, China, Germany, Iran, the Netherlands, Portugal, South Korea, the UK, or CA, MI, NJ states. The graph-only representations for Australia and Belgium appeared to indicate an excess of men (**Figure 5C-E**). For the other 10 countries, the proportion of men among deaths ranged from 55% (Norway) to 68% (Italy) (**Table 3**).

**Figure 5:**
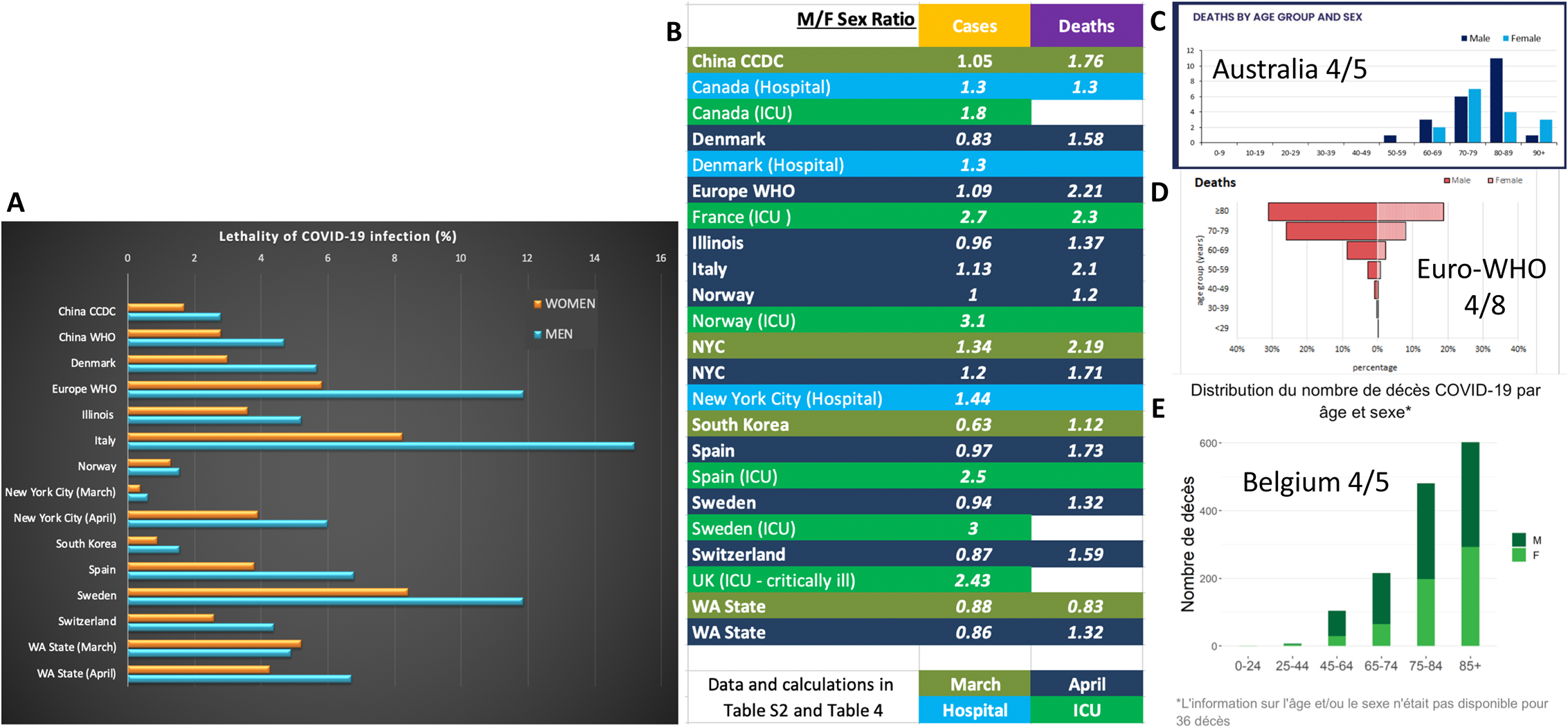
Sexual dimorphism in lethality of COVID-19 infection. **A**: Lethality (or case fatality, calculated as the percentage of deaths among confirmed cases) by sex for data sets where disaggregated number of cases and deaths was found for the same cohort. Data source and calculations shown in Table S2. **B:** Calculated sex ratios (number of men/number of women) for cases and deaths quantify the excess of men among deaths and ICU cases. **C:** Three entities provided graphic visualization of deaths by sex and age. Age binning, type of graph, and metrics were different in all three. No numbers were provided for Belgium or Australia, and we were unable to calculate a sex ratio.

#### Calculated metrics - Lethality and Sex Ratio

When data were available for both deaths and cases on the same cohort, we systematically calculated the lethality for each sex (fraction of deaths per confirmed cases, in men and in women, shown in **Fig. 5A**) and the excess of men (male-to-female sex ratio) among cases and deaths (**Fig. 5B**). (Data and calculations shown in **Supplementary Table S2** or **Table 4**).

**Table 4:** Sex ratio among hospitalizations and ICU cases. Lethality (shown in red font) was calculated as number of deaths/number of cases for each sex. Percentage was calculated as number of men/number of women in same category. Italics indicate values calculated by the authors. Cells highlighted in pale green indicate metrics displayed on websites. UK data: ref. 30; other data sources shown in Table 1.

|  |  |  | Number in<br>report with<br>known sex | Number |  | Percentage |  |
| --- | --- | --- | --- | --- | --- | --- | --- |
| Date of Data |  |  |  | Women | Men | Women | Men |
| Canada (Hospital) | April 9 | Cases | 1479 | 641 | 838 | 43 | 57 |
|  |  | Deaths | 278 | 125 | 163 | 45 | 55 |
|  |  | *Lethality* |  | *19.50%* | *19.40%* |  |  |
| Denmark (Hospital) | April 9 | Cases | 1,444 | 629 | 815 | 43.6 | 56.4 |
| New York City (Hospital) | April 19 | Cases | 33,234 | 13,594 | 19,640 | 40.9 | 59.1 |
| Canada (ICU) | April 9 | Cases | 408 | 145 | 263 | 36 | 64 |
| France (ICU ) | 3/16-4/5 | Cases | 2,218 | 599 | 1619 | 27 | 73 |
|  |  | Deaths | 187 | 56 | 131 | 30 | 70 |
|  |  | *Lethality* |  | *9.30%* | *8.10%* |  |  |
| Norway (ICU) | April 9 | Cases | 174 | 42 | 132 | 24 | 76 |
| Spain (ICU) | April 6 | Cases | 3,078 | 884 | 2,194 | 28.7 | 71.3 |
| Sweden (ICU) | April 15 | Cases | 954 | 241 | 713 | 25.3 | 74.7 |
| UK (ICU - critically ill) | Mar 20 | Cases | 196 | 57 | 139 | 29.1 | 70.9 |

An apparent excess of men amongst deaths was evidenced by a sex ratio ranging from 1.2 (20% more men) in Norway to 2.1 (more than twice as many men) in Italy in April data. Also, in April data, all countries and states also showed higher lethality in men compared to women, irrespective of the number or sex ratio in cases or average lethality in the country.

Lethality varied greatly from over 15% in Italian men (almost twice as high as in Italian women) to under 2% in Norway (which showed the smallest disparity between men and women lethality). Country-specific policies, infrastructure, climate and/or lifestyle may underlie the puzzling differences in lethality. Denmark, with a similar number of cases as Norway (both in absolute numbers and per capita, 1,329 and 1, 326/1M population respectively; Worldometers.com 4/21/20 data) had a lethality three times higher, with a strong male bias. Sweden, with a slightly higher per capita COVID-19 infection rate (1,517) recorded the second largest lethality at almost 12% in men.

Switzerland, which has the third highest per capita infection rate among countries with at least 5 million inhabitants (3,243/1M; behind only Spain and Belgium and ahead of Italy), had the same sex ratio in cases (~0.87) and deaths (~1.59) as Denmark, but a much lower lethality, in both sexes. Lethality increased dramatically in New York City as the epidemic progressed (from <1% to 3.9% for women and 6% for men), but the sex difference was maintained. This is similar to what was observed in China where the lethality had doubled between the two February reports, but the difference of lethality between the sexes remained similar. Washington state, which was the first recognized site of COVID-19 outbreak in the US, already had a lethality around 5% in late March, for both sexes (the only such finding). By April, average lethality was unchanged, but the sex difference had emerged.

### Severity of disease: ICU and hospitalizations

A March 20 report from the Intensive Care National Audit and Research Centre (inarc.org) disclosed that, of the 196 critically ill patients in the UK, 70.9% were male^30^. None of the other reported characteristics of the cohort (including Body Mass Index, comorbidities, length of stay, deaths, and therapies) were disaggregated by sex.

In-hospital observation days in the ChinaCDC report was 342,063 for men and 319,546 for women (Table 1 in^1^). This metric has typically been used as a proxy for severity but, in the case of a lethal infectious disease, there can be ambiguity as to whether observation stops because the patient has died or recovered. Additionally, numbers could be confounded by cultural factors (*e.g*. resulting in one sex or the other being admitted later). In any case, in this cohort, we were able to calculate that the average number of observation days per patient was similar in both sexes (342,063 days/22981 = 14.88 cases for men; 319,546/21,691 = 14.73 for women).

Where sex-disaggregated numbers were available (**Table 4**), data showed that men represented the vast majority of cases admitted in the ICU, with percentages ranging from 64% in Canada to 76% in Norway. (All the countries for which the information was known had either roughly similar number of women and men or a higher proportion of women among the total cases). A similar trend, with a lower sex ratio divergence, was seen among hospital admissions, where women represented 41-45%. (An analysis of 1,099 hospitalized patients in China, published in the New England Journal of Medicine (NEJM), reported 41.9% of women^31^).

Two countries provided numbers of cases and deaths on the same cohorts: Canada among severe hospitalized cases, and France among ICU cases. For these we calculated lethality for each sex (Table 4, in red). In striking contrast to lethality in the infected population at large, we found similar (19.5% in Canada) or even slightly higher lethality in women than men (France, 9.3% vs. 8.1%) among those severe cases. This suggests that while women may be developing a less severe (or different, see below) form of disease, those who do reach the ICU in deep respiratory distress may have a similarly poor chance of survival as men.

### Comorbidities, symptoms

The US National Center for Health Statistics reported that, among 1,879 deaths with pneumonia <u>and</u> COVID-19 reported as of 4/4/20, 1,097 (58.4%) were men; in contrast, only 52% of deaths with pneumonia without COVID-19 were men, and similar numbers of men and women had died from influenza in the same period (2,214 men vs. 2,253 women; 49.6% men).

Spain was the only country for which we could find a detailed report of comorbidities by sex (data April 6; data captures and sources for US and Spain shown in **Supplementary Figure S1**). Numbers and percentages were reported for 11 different symptoms and 4 comorbidities, and p-values were provided. While it was unclear to what those p-values and some of the percentages referred, numerical data indicated that sore throat, vomiting, and diarrhea were more frequent in women. The widely described symptoms of COVID-19 appear to be those most frequently found in men than women: fever, pneumonia, severe acute respiratory distress syndrome and other respiratory symptoms. Pneumonia, for example, was found in 65% of men, but only 49% of women.

## Discussion

### Sex-disaggregated data available to understand the characteristics of COVID-19 are limited

Our study showed that sex-disaggregating reported data is still not a widespread practice. For example, Italy has, since the beginning of the crisis in this country, maintained a daily updated, publicly available GitHub database documenting cases, deaths, number of tests performed, intensive care patients, and recovery rates, but no sex information. The strong disparity of lethality between men and women highlighted in the February reports from China likely prompted more countries to start reporting metrics by sex over time. However, even by mid-April, no data was found for many countries or US states, even on number of confirmed cases.

Beyond infection and death rates, we found gendered information about tests performed for one US State (Illinois), comorbidities for one country (Spain), and therapies provided, recovery rates, or possible method of transmission for none.

It is unclear whether the information is collected but not widely released or analyzed, or if it is not collected at all. Whatever data existed early on seemed to have been available only to the institutions that collected them, and released in priority to news outlets (including scientific journals). While this allowed a joint report by CNN and the British Medical Journal (BMJ) (in blog format, prior to peer review) of early sex-disaggregated trends (published after we had performed the first data capture), it highlighted an inequality in accessibility to data. The urgency of the developing epidemic has resulted in most of the available data and analyses being published in the form of press releases, blogs, Tweets, or non-peer-reviewed pre-prints. Accessibility was limited for scholars worldwide, and notably even the powerful news media outlet CNN has published that the US CDC did not respond to a request for sex-disaggregated data for their study^7^.

### Sex-disaggregated metrics available to understand the characteristics of the disease are disparate

When available, the information was presented in a wide variety of ways, strikingly illustrated in Fig. 2. More importantly, we found extreme diversity in reported metrics: number, rate per 100,000 population, fraction of various variables, percentage of men, percentage of women, etc. When possible, we used available data to reverse calculate and present complete data sets with comparable metrics. This was time-consuming, error-prone, and sometimes not possible. For example, while reporting cases per 100,000 population is very valuable in epidemiology (to calculate mortality rates at the population level), calculating actual numbers from this metric requires knowing the total population size at the time of data collection, which is typically not provided.

Similarly, for sex- and age-disaggregated data, systematic comparison was not possible because of the variety of age range options chosen by the various entities to bin their data. It is unclear if age information was collected as a number, which was later binned during analysis, or if questionnaires collecting data already limited answers to age brackets. If the latter, even access to raw data will not allow future comparison between data sets.

To fully understand if the higher proportions of men or women among subcohorts represent a true excess of one sex, values need to be related to baseline population numbers for each subset. The overlay of the national age pyramid on the bar graph for Spain data (Fig. 2) was helpful in this respect. It, however, highlighted further limitations, as the data used for the baseline population was almost a year old (and no numerical values were made available).

### Standardization of data collection and data points needs to be improved

While all methods of graphing and metrics may be valuable for different purposes, comparable, standardized raw data need to be accessible to scholars for meaningful, statistically significant analysis. For continuous variables *(e.g*. age), actual numerical values are needed. We could not find any stated justification on any governmental website for the wide array of age-binning categories illustrated in Fig. 2. Not only does it make comparison between data sets difficult, it might hide - or artificially create - trends. For example, grouping children aged 10-19, across puberty, is likely to mask hormonally influenced outcomes. Rigorous statistical analysis needs to be applied to raw data to reveal trends (and guide age-binning), rather than the converse.

For sex, provision for a third option, beyond the binary, would be useful to accommodate both various country-specific laws and missing data. When this was provided, entities calculated proportions of women/men either out of the total (leaving percentages not adding to 100%, as in WA state) or out of the cases for which sex was known (resulting in information being reported on a limited fraction of total cases - see legend of Table 3).

Where countries have put online high-quality disaggregated data (*e.g*. Canada, https://www150.statcan.gc.ca/t1/tbl1/en/tv.action?pid=1310076601), those are typically coded (*e.g*. 1 =male, 2=female). This is, of course, useful for automated analysis of data. It does, however, increase potential for data entry error and makes it difficult for a human eye to perform quality control on the data sets. We suggest entering the data in a standardized (*e.g*. each country will collect male, female, other) but non-coded way, sensitive to local usages, in the local language. Software can then be designed to code the entry (*i.e*. transform woman, femme, kvinna, donna, mujer, sieviete, babae, or Frau into “2”), merge the standardized data sets, and redeploy them in the native language of the country for easy use by the local researchers.

We recommend transparency and real-time data release, both to allow timely scholarly analysis and to limit potentially politically motivated retention of data by governments. Increased international preparedness will also be required if collected data are to provide accurate sources to guide scientific discovery and policy decision-making. This will require enhanced international cooperation between epidemiologists, clinicians, statisticians, biologists, ethicists, and local, federal, governmental and trans-national entities, to agree on standardized metrics, and stable data storage systems, to collect relevant information while ensuring the privacy of citizens.

### Increase of proportion of women cases over time

The Danish data (shown in the mirror bar graphs of cases disaggregated by both sex and age in Fig. 3 and in comparison with other countries in Fig. 4) illustrate both the trend and the difficulty (futility?) of interpreting daily updated data in the exponential growth phase of an epidemics. Policies based on the early trend showing a massive apparent excess of men at all ages would have been rendered obsolete just 3 weeks later.

This increase in the proportion of women during the course of the epidemic was seen in all surveyed regions (except for South Korea, which already reported a large excess of women among confirmed cases by the time of our first data capture).

Several hypotheses can be made, all of which require further, currently unavailable sex-disaggregated data to prove or disprove. It is possible that more accurate statistics were obtained with increasing cohort size or that higher lethality in men started reducing the sex ratio of cases as the epidemic progressed. It is also possible that expansion of testing availability resulted in milder cases, or cases with different presentation, becoming counted in women. It could also be a true increase in female rate of infection over time, possibly due to evolution of exposure type and different societal gender roles. For example, in societies where a smaller fraction of women work outside of the home, infection of women may become more widespread in a second wave after infected men have brought the disease back to their communities and their caretakers. Granular sex-disaggregated data about likely method of transmission or disease symptoms will be necessary to test these hypotheses.

### Higher severity of the disease in men

A joint CNN/BMJ press release reported that “men were 50% more likely than women to die after being diagnosed with COVID-19^7^”. Our analysis recapitulated this trend, but more specific data will be needed for scientists to understand the underlying causes of the findings, as well as the exceptions.

The similar lethality rates for men and women in ICU in France or severe hospitalized cases in Canada (Table 4), if generalizable, suggest a more nuanced picture. Additionally, if women are offered testing at a lower rate, they may be dying as well, but not be included in the counts because they have not been formally diagnosed. If the presentation of the disease is different in men and women, the criteria for testing, established on the earlier, male-biased statistics, may be inadequate to identify the infected female population. There may also be regional, cultural, and infrastructural specificities resulting in differential reporting of outcomes for men and women. For example, several anecdotal reports (*e.g*. from Italy) suggested deaths may be underreported in real time, and that women may be dying at home rather than in the hospital in greater numbers, and thus be underreported by overwhelmed local authorities.

### Further research is needed on mechanisms underlying sex differences in COVID-19 severity and infection rates

While the rate of death has been reported to be higher when infection is associated with pre-existing conditions such as with cardiovascular disease, diabetes, lung disease, or cancer, it is unclear if any one of these comorbidities have a disproportionate effect in one sex or the other because these data have yet to be widely reported by sex. Both the US CDC and Spanish data suggest that the severe lung disease and pneumonia is a feature more frequently associated with COVID-19 in men. The Spanish data also indicate that other symptoms may have a sexually dimorphic prevalence, with diarrhea, vomiting or sore throat being part of the clinical symptoms more frequently observed in women. This suggests that the extent of infection in women may not have been fully recognized because presentation may be different. Perhaps the large press coverage given to by the most famous celebrity couple known to have survived COVID-19, American actors Tom Hanks and Rita Wilson, who shared that they experienced very different symptoms during their joint quarantine, will prod countries to document those in a sex-disaggregated way in the future.

To explain the higher lethality in men and the higher rate of smokers among non-survivors (9%) than survivors (5%) in a small (191 patients) study^32^ much has been made of the fact that, in China, smoking is reported to be a widely sexually dimorphic activity, where ~50% of the men, but only 2% of women, smoke 40% of the world’s tobacco (e.g.^6,33^. However, divergent information has emerged. In the NEJM report of a cohort of hospitalized patients in China, 78% of even severe cases were never smokers^31^. In France smoking rates are more similar in men and women (28.2% in men and 22.9% of women in 2018). Yet, only 8.5% of 11,000 people admitted to all Paris hospitals with COVID-19 as of early April were smokers^34^. An April 21 preprint article examining the proportion of daily smokers among a single-center cohort of 343 inpatients and 149 outpatients in Paris, reported that only about 5% of the infected were smokers, in both sexes^35^. While this study has yet to be peer-reviewed and reproduced in other settings, it again highlights the broader need for such systematic data collection and analysis by sex.

Literature studying biological mechanisms underlying sex differences in COVID-19 is still nonexistent at this early time. We found one preprint in which the authors designed *in silico* experiments in search of a possible mechanistic explanation for the sex differences in SARS-CoV-2 infection^36^. Using publicly available human tissue gene expression and ChIP-Seq data sets, they proposed an intriguing hypothesis where the androgen receptor, AR could directly control the expression of ACE2, one of the two main proteins for SARS-CoV-2 entry into human cells (the other, TMPRSS2 is a known androgen-responsive gene^37^). Their analysis of single-cell RNA-sequencing found that more pulmonary alveolar type II cells express *ACE2* in men than women and that *ACE2* is expressed in the prostate and in Sertoli and Leydig cells of the testis, possibly providing another entry path for the virus in men. Expression of *AR* and *ACE2* correlated positively in all tissue types and ChIP-Seq datasets identified a putative AR-binding site upstream of the ACE gene. If these *in silico* explorations withstand the rigors of peer-review and of *in vitro* or *in vivo* experiments, it may mean that aspects of COVID-19 pathology will be susceptible to anti-androgen therapy (as is used routinely in prostate cancer).

### Overall conclusions

Through capture, collation, and systematic analysis of datasets from 20 countries and 6 US states, we found wide-ranging disparity in the format, sex- and/or age-disaggregation, and amount and accessibility of data made available to the public and scholars. Data on confirmed COVID-19 cases and outcomes stratified by sex was unavailable for most countries and US states surveyed. This paucity of accessible raw datasets and disparate metrics used to capture data make it difficult to draw comparisons and meaningful conclusions to inform healthcare, public health, and policy strategies.

In an infectious disease outbreak, different geographic areas are at different stages of the epidemic at different times, and the data themselves evolve quickly as the events unfold. Even standardized data will be difficult to compare and interpret for policy-making. The ascending curve of a pandemic is obviously not the best time to start devising strategies and building infrastructure to collect global standardized data for efficient surveillance. Throughout the evolution of the COVID-19 epidemic, we noted improved reporting as most countries and US states strove to make more data available. Countries had clearly disparate resources available for this enterprise (access to testing, reliable public health institutions, and informatic platforms), and preparedness might have helped alleviate the reporting burden for lower-resource regions.

Lack of data harmonization and sex-disaggregation, however, were ubiquitous issues. The US CDC’s COVID-19 Case Report Form^38^ does collect, for each Person Under Investigation (PUI), sex, symptoms, pre-existing conditions, dates of hospitalization and treatment received, potential source of infection, and testing status. It is critical that *Sex* as a *Biological Variable* be considered an essential metric, rather than an afterthought, as we improve international efforts toward capture, and accessibility, of standardized data.

## Data Availability

This systematic analysis used only publicly available data. A complete list of sources used has been collated in Table 1 of this manuscript.

## Acknowledgements

The authors thank Yulong Fu, Lisa Guay-Woodford, Susan Knoblach, Hiroki Morizono, Eric Vilain, and Maygon Wendel for helpful suggestions during the conception of this report.

## Supplementary Material

**Figure S1:**
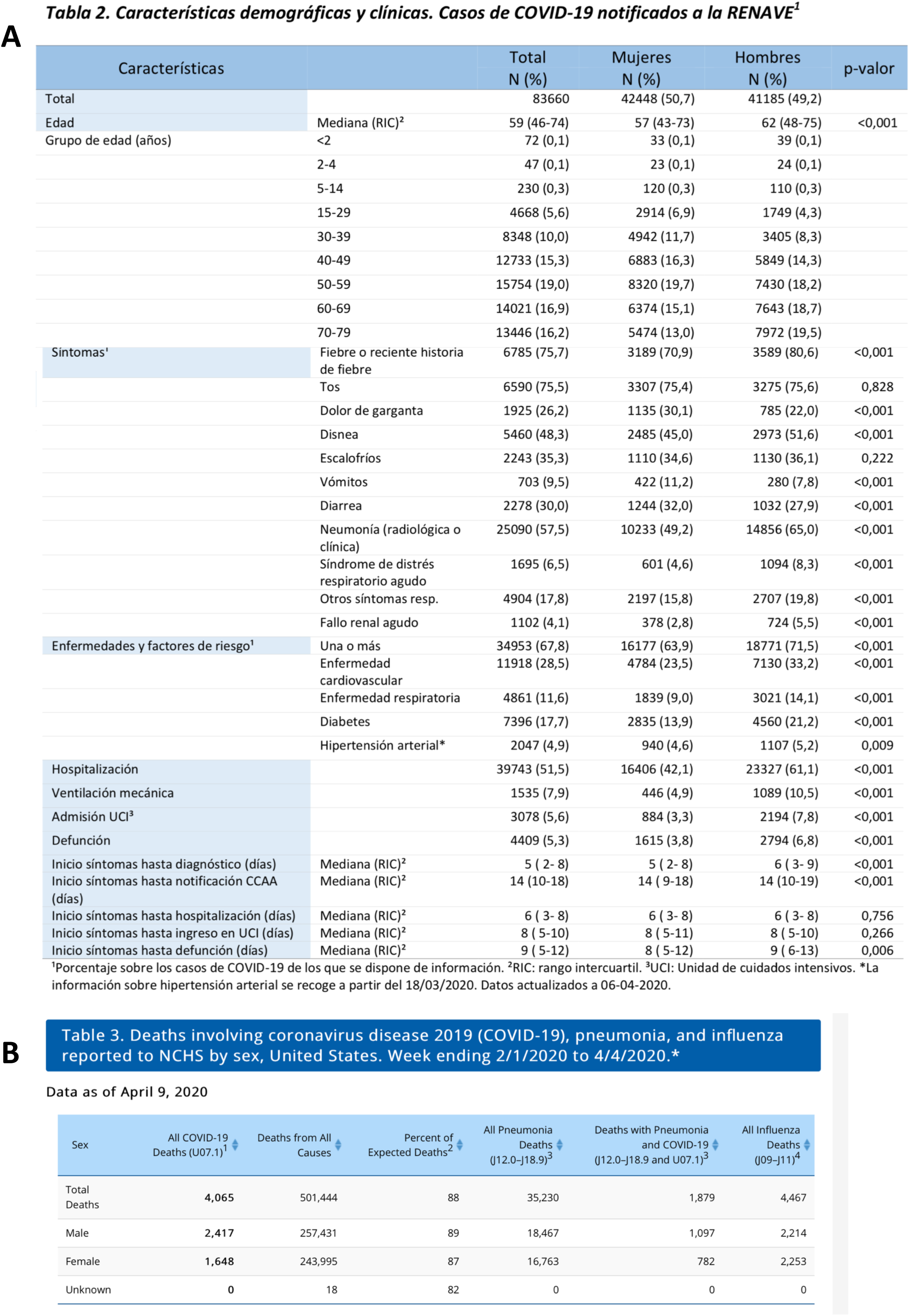
Screen captures of sex-disaggregated data of comorbidities and symptoms of COVID-19 infection. **A**: in the Spanish population as of 4/6/20, accessed 4/9/20 on the following URL: https://www.isciii.es/QueHacemos/Servicios/VigilanciaSaludPublicaRENAVE/EnfermedadesTransmisibles/Documents/INFORMES/Informes%20COVID-19/Informe%20n°%2021.%20Situación%20de%20COVID-19%20en%20España%20a%206%20de%20abril%20de%202020.pdf **B**: in the US population, for the 2/1–4/4/2020 period, accessed 4/10/20 at the following URL: https://www.cdc.gov/nchs/nvss/vsrr/COVID19/index.htm

**Table S1:** Confirmed cases of COVID-19 infection by sex and age (raw data and calculations for Fig. 4) All cases over age 80 were combined for consistency among datasets. The dataset from Spain was originally age-binned as follows: <2, 2–4, 5–14, 15–29, 30–39, 40–49, 50–59, 60–69, 70–79, >80. In order to visualize Spain dataset graphically with other countries, raw case numbers for groups “<2,” “2–4,” and “5–14” were combined into one group labeled “<15.” The rest of the Spain dataset age-bins (15–30, 30–40, 40–50, 50–60, 70–80, and >80) remain the same. Italics denote calculations performed on the raw data by the authors.

|  | Belgium (Apr 5) |  |  |  | Denmark (Mar 22) |  |  |  | Denmark (Apr 7) |  |  |  | Italy (Apr 7) |  |  |  | Norway (Apr 9) |  |  |  | Switzerland (Apr 9) |  |  |  |
| --- | --- | --- | --- | --- | --- | --- | --- | --- | --- | --- | --- | --- | --- | --- | --- | --- | --- | --- | --- | --- | --- | --- | --- | --- |
| Age Range | Female | Male | Combined | Male Cases (%) | Female | Male | Combined | Male Cases (%) | Female | Male | Combined | Male Cases (%) | Female | Male | Combined | Male Cases (%) | Female | Male | Combined | Male Cases (%) | Female | Male | Combined | Male Cases (%) |
| <10 | 55 | 71 | 126 | 56.3 | 11 | 2 | 13 | 15.4 | 27 | 31 | 58 | 53.4 | 375 | 450 | 825 | 54.5 | 37 | 31 | 68 | 45.6 | 7 | 5 | 12 | 41.7 |
| 10-20 | 107 | 87 | 194 | 44.8 | 21 | 14 | 35 | 40 | 70 | 59 | 129 | 45.7 | 596 | 621 | 1217 | 51.0 | 136 | 121 | 257 | 47.1 | 27 | 24 | 51 | 47.1 |
| 20-30 | 1114 | 507 | 1621 | 31.3 | 77 | 88 | 165 | 53.3 | 388 | 219 | 607 | 36.1 | 3181 | 2426 | 5607 | 43.3 | 517 | 354 | 871 | 40.6 | 130 | 91 | 221 | 41.2 |
| 30-40 | 1419 | 733 | 2152 | 34 | 81 | 110 | 191 | 57.6 | 451 | 294 | 745 | 39.5 | 4780 | 4158 | 8938 | 46.5 | 488 | 496 | 984 | 50.4 | 171 | 135 | 306 | 44.1 |
| 40-50 | 1797 | 1180 | 2977 | 39.6 | 126 | 249 | 375 | 66.4 | 609 | 496 | 1105 | 44.9 | 8701 | 7444 | 16145 | 46.1 | 565 | 591 | 1156 | 51.1 | 230 | 181 | 411 | 44.0 |
| 50-60 | 1883 | 1694 | 3577 | 47.4 | 86 | 162 | 248 | 65.3 | 626 | 499 | 1125 | 44.4 | 11608 | 12648 | 24256 | 52.1 | 563 | 666 | 1229 | 54.2 | 275 | 316 | 591 | 53.5 |
| 60-70 | 1053 | 1565 | 2618 | 59.8 | 55 | 93 | 148 | 62.8 | 345 | 393 | 738 | 53.3 | 7306 | 13057 | 20363 | 64.1 | 338 | 392 | 730 | 53.7 | 141 | 226 | 367 | 61.6 |
| 70-80 | 1095 | 1443 | 2538 | 56.9 | 48 | 70 | 118 | 59.3 | 258 | 349 | 607 | 57.5 | 7990 | 13581 | 21571 | 63.0 | 227 | 264 | 491 | 53.8 | 184 | 218 | 402 | 54.2 |
| >80 | 2158 | 1628 | 3786 | 43.0 | 42 | 60 | 102 | 58.8 | 302 | 219 | 521 | 42.0 | 13549 | 11519 | 25068 | 46.0 | 211 | 163 | 374 | 43.6 | 339 | 204 | 543 | 37.6 |
| Total | 10681 | 8908 | 19589 | - | 547 | 848 | 1395 | - | 3076 | 2559 | 5635 | - | 58086 | 65904 | 123990 | - | 3082 | 3078 | 6160 | - | 1504 | 1400 | 2904 | - |

|  | Spain (Apr 6) |  |  |  |
| --- | --- | --- | --- | --- |
| Age Range | Female | Male | Combined | Male Cases (%) |
| <15 | 176 | 173 | 349 | 49.6 |
| 15-30 | 2914 | 1749 | 4663 | 37.5 |
| 30-40 | 4942 | 3405 | 8347 | 40.8 |
| 40-50 | 6883 | 5849 | 12732 | 45.9 |
| 50-60 | 8320 | 7430 | 15750 | 47.2 |
| 60-70 | 6374 | 7643 | 14017 | 54.5 |
| 70-80 | 5474 | 7972 | 13446 | 59.3 |
| >80 | 7122 | 6687 | 13809 | 48.4 |
| Total | 42205 | 40908 | 83113 | - |

**Table S2:** Data and calculations of lethality and sex ratio for entities providing sex-disaggregated data for cases and deaths on the same cohort. Lethality was calculated as the percentage of deaths among cases. Sex ratio was calculated as the fraction of number of men over number of cases. Rows for early captures are highlighted in **olive green**, late captures in **dark blue**. Italics indicate calculations by the authors, pale sage green cells indicate metrics documented on the websites. (China data from refs. 1 & 2; South Korea data from ref. 29.)

|  |  |  | Number<br>in report<br>with<br>known<br>sex | Number |  | Percentage |  |  |
| --- | --- | --- | --- | --- | --- | --- | --- | --- |
| Date of Data |  |  |  | Women | Men | Women | Men | Sex Ratio |
| China CCDC | Feb 11 | Cases | 44,672 | 21,691 | 22,981 | 48.6 | 51.4 | 1.05 |
|  |  | Deaths | 1,023 | 370 | 653 | 36.2 | 63.8 | 1.76 |
|  |  | Lethality (%) |  | 1.7 | 2.8 |  |  |  |
| China WHO | Feb 20 | Lethality (%) |  | 2.8 | 4.7 |  |  |  |
| South Korea | Mar 21 | Cases | 8,799 | 5412 | 3387 | 61.5 | 38.5 | 0.63 |
|  |  | Deaths | 102 | 48 | 54 | 47.1 | 52.9 | 1.12 |
|  |  | Lethality (%) |  | 0.89 | 1.56 |  |  |  |
| New York City | Mar 22 | Cases | 12,322 | 5,255 | 7,067 | 43 | 57 | 1.34 |
|  |  | Deaths | 63 | 20 | 43 | 31.8 | 68.2 | 2.19 |
|  |  | Lethality (%) |  | 0.38 | 0.61 |  |  |  |
| WA State | Mar 23 | Cases | 2,132 | 1,133 | 999 | 53 | 47 | 0.88 |
|  |  | Deaths | 108 | 59 | 49 | 55 | 45 | 0.83 |
|  |  | Lethality (%) |  | 5.2 | 4.9 |  |  |  |
| Denmark | Apr 9 | Cases | 5,635 | 3,076 | 2,559 | 55 | 45 | 0.83 |
|  |  | Deaths | 237 | 92 | 145 | 38.8 | 61.2 | 1.58 |
|  |  | Lethality (%) |  | 2.99 | 5.67 |  |  |  |
| Europe WHO Region | Apr 8 | Cases | 95,401 | 45,660 | 49,741 | 48 | 52 | 1.09 |
|  |  | Deaths | 8,569 | 2,666 | 5,903 | 31 | 69 | 2.21 |
|  |  | Lethality (%) |  | 5.83 | 11.87 |  |  |  |
| Illinois | Apr 19 | Cases | 28,713 | 14,645 | 14,068 | 51 | 49 | 0.96 |
|  |  | Deaths | 1,254 | 529 | 725 | 42.2 | 57.8 | 1.37 |
|  |  | Lethality (%) |  | 3.6 | 5.2 |  |  |  |
| Italy | Apr 7 | Cases | 124,162* | 58,172 | 65,990 | 46.9 | 53.1 | 1.13 |
|  |  | Deaths | 14,840 | 4,793 | 10,047 | 32.3 | 67.7 | 2.1 |
|  |  | Lethality (%) |  | 8.23 | 15.2 |  |  |  |
| Norway | Apr 9 | Cases | 6,160 | 3,082 | 3,078 | 50 | 50 | 1 |
|  |  | Deaths | 88 | 40 | 48 | 45 | 55 | 1.2 |
|  |  | Lethality (%) |  | 1.30 | 1.56 |  |  |  |
| New York City | Apr 19 | Cases | 126,098 | 59,484 | 66,614 | 47 | 53 | 1.2 |
|  |  | Deaths | 6,331 | 2,337 | 3,994 | 36.9 | 63.1 | 1.71 |
|  |  | Lethality (%) |  | 3.9 | 6 |  |  |  |
| Spain | Apr 6 | Cases | 83,633* | 42,448 | 41,185 | 50.8 | 49.2 | 0.97 |
|  |  | Deaths | 4,409 | 1,615 | 2,794 | 36.7 | 63.3 | 1.73 |
|  |  | Lethality (%) |  | 3.8 | 6.78 |  |  |  |
| Sweden | Apr 15 | Cases | 11,927 | 6,143 | 5,784 | 51.5 | 48.5 | 0.94 |
|  |  | Deaths | 1,203 | 517 | 686 | 43 | 57 | 1.32 |
|  |  | Lethality (%) |  | 8.4 | 11.86 |  |  |  |
| Switzerland | Apr 9 | Cases | 24,140 | 12,876 | 11,264 | 53.3 | 46.7 | 0.87 |
|  |  | Deaths | 804 | 311 | 493 | 38.7 | 61.3 | 1.59 |
|  |  | Lethality (%) |  | 2.6 | 4.38 |  |  |  |
| WA State | Apr 19 | Cases | 11,448 | 6,137 | 5,311 | 53 | 47 | 0.86 |
|  |  | Deaths | 624 | 268 | 356 | 43 | 57 | 1.32 |
|  |  | Lethality (%) |  | 4.37 | 6.7 |  |  |  |

